# Longitudinal neonatal brain development and socio-demographic correlates of infant outcomes following preterm birth

**DOI:** 10.1101/2022.08.11.22278469

**Authors:** Lucy Vanes, Sunniva Fenn-Moltu, Laila Hadaya, Sean Fitzgibbon, Lucilio Cordero-Grande, Anthony Price, Andrew Chew, Shona Falconer, Tomoki Arichi, Serena J. Counsell, Joseph V. Hajnal, Dafnis Batalle, A. David Edwards, Chiara Nosarti

## Abstract

Preterm birth results in premature exposure of the brain to the extrauterine environment during a critical period of neurodevelopment. Consequently, infants born preterm are at a heightened risk of adverse behavioural outcomes in later life. We characterise longitudinal development of neonatal regional brain volume and functional connectivity in the first weeks following preterm birth, sociodemographic factors, and their respective relationships to psychomotor outcomes and psychopathology in toddlerhood. We study 121 preterm infants preterm who underwent magnetic resonance imaging shortly after birth, at term-equivalent age, or both. Longitudinal regional brain volume and functional connectivity were modelled as a function of psychopathology and psychomotor outcomes at 18 months. Better psychomotor functioning in toddlerhood was associated with greater relative right cerebellar volume and a more rapid decrease over time of sensorimotor degree centrality in the neonatal period. In contrast, increased 18-month psychopathology was associated with a more rapid decrease in relative regional subcortical volume. Furthermore, while socio-economic deprivation was related to both psychopathology and psychomotor outcomes, cognitively stimulating parenting predicted psychopathology only. Our study highlights the importance of longitudinal imaging to better predict toddler outcomes following preterm birth, as well as disparate environmental influences on separable facets of behavioural development in this population.

## 1. Introduction

Individuals born preterm (i.e., before 37 completed weeks of gestation) are at an increased risk of experiencing psychomotor delay and behavioural problems in early development (Allotey et al., 2018; Johnson and Marlow, 2011), which can be predictive of cognitive and socio-emotional outcomes in later life (Arpi and Ferrari, 2013; Briggs-Gowan and Carter, 2008; Johnson and Marlow, 2014). Although the severity of adverse developmental outcomes appears to show a gestational age related gradient (Johnson, 2007; Moore et al., 2013), even those born moderately-to-late preterm have a significantly higher risk of non-optimal outcomes compared to those born at full term (Cheong et al., 2017; Johnson et al., 2018; Kerstjens et al., 2011; Woythaler, 2019). Overall, the developmental consequences of preterm birth vary widely, yet the early predictors and moderators of behavioural outcomes remain poorly understood. Alterations in brain development as a result of premature exposure of the brain to the extrauterine environment during a critical period of neural growth is thought to play a substantial role in conferring risk for developmental problems (Ment et al., 2009); however, it is also clear that the environment can have a significant impact on behavioural development in this vulnerable population (Neel et al., 2018; Vanes et al., 2021; Wong and Edwards, 2013).

Psychomotor functioning (including cognitive, language, and motor skills) and psychopathology (including emotional and behavioural problems) are intimately connected throughout development. For example, infants exhibiting significant psychomotor delay are more likely to develop behavioural problems in later childhood (Caplan et al., 2015; Friedlander et al., 1982). At the same time, preterm infants are at an increased risk of psychopathology even after controlling for concomitant psychomotor delay (Delobel-Ayoub et al., 2006; Treyvaud et al., 2012). Neuroimaging research in preterm cohorts, however, frequently focuses on these facets of behaviour in isolation, thus potentially failing to disentangle the unique mechanisms contributing to psychopathology and psychomotor functioning during early development. Furthermore, studies probing associations between neonatal brain development and infant outcome typically include imaging at only one timepoint in the neonatal period; however, it is possible that trajectories of brain development more accurately reflect the effect of pathology on the establishment of neural systems and are thus more predictive of behavioural outcome than cross-sectional metrics (Duerden et al., 2013; Hazlett et al., 2017; Young et al., 2017).

Neonatal imaging of the preterm brain at term-equivalent age has been shown to be variably predictive of developmental outcomes (Kwon et al., 2014). Research investigating the neonatal neural correlates of psychopathology has mainly focused on specific characteristics pertinent to the so-called preterm behavioural phenotype, encompassing anxiety, socio-emotional, and attention-related problems in early childhood (Brenner et al., 2021b). For example, socio-emotional problems have been associated with reduced regional brain volumes (Rogers et al., 2012), alterations in white matter connectivity (Brenner et al., 2021a; Kanel et al., 2021; Rogers et al., 2016) and functional connectivity (Kanel et al., 2022; Rogers et al., 2017; Sylvester et al., 2018) in the neonatal period. These effects, however, also show considerable overlap with the potential early neural mechanisms implicated in cognitive, motor, and language development, such as subcortical volume reductions (Chau et al., 2019; Loh et al., 2017), structural and functional connectivity alterations in sensory and cognitive networks (Ball et al., 2015; Gozdas et al., 2018; Peyton et al., 2020; Rogers et al., 2017). Therefore, studying the neural correlates of early risk for psychopathology and psychomotor problems within the same cohort – each while controlling for the other – will help disentangle the unique mechanisms underlying these conceptually separable domains.

In addition, it is important to take into account environmental and familial factors which are known to impact early development, both on a behavioural and on a neural level. Factors such as socio-economic disadvantage may predispose individuals to altered trajectories of brain development, with significant implications for mental health. Neonatal whole-brain connectivity of several subcortical and cortical regions for example has been shown to mediate the relationship between socio-economic status and behavioural inhibition and externalising problems at 2 years of age in the general population (Ramphal et al., 2020). Moreover, evidence also suggests that favourable socio-economic status can mitigate the effects of early brain injury on behavioural outcomes following preterm birth (Benavente-Fernández et al., 2019). Socio-economic factors may furthermore confound the relationship between psychopathology and neurocognitive outcomes in preterm children (Lowe et al., 2019). In addition, the more immediate home environment plays a crucial role in child development, with family factors being particularly important in differentiating resilient and behaviourally vulnerable preterm-born children (Lean et al., 2020) and cognitively stimulating parenting showing a specific association with reduced psychopathology in this population (Vanes et al., 2021).

In this study, we investigate both the structural and functional neonatal neural correlates of developmental outcomes following preterm birth – taking into account both psychomotor development and behavioural indicators of psychopathology. Using data from the Developing Human Connectome Project (Edwards et al., 2022), we leverage longitudinal imaging in the neonatal period (immediately following preterm birth and at term-equivalent age) in order to test whether the *rate* of development in brain structure or function is associated with outcome in toddlerhood, while controlling for key environmental socio-demographic factors (socio-economic deprivation and cognitively stimulating parenting). We use voxelwise measures of local volume and whole-brain functional connectivity, allowing us to test for regionally specific developmental effects associated with behaviour. We also separately investigate associations between environmental factors and developmental outcomes.

## 2. Methods

### 2.1. Participants

205 infants born preterm (<37 weeks gestation) underwent at least one MRI session in the neonatal period as part of the Developing Human Connectome Project (dHCP, http://www.developingconnectome.org/, 3^rd^ release; Edwards et al., 2022). Ethical approval was given by the London - Riverside Research Ethics committee (REC ref: 14/LO/1169), and written consent was obtained from all participating families prior to imaging. Infants were imaged either shortly after birth, at term-equivalent age, or both (N = 64). Preterm-born infants were recruited from the neonatal unit and postnatal wards at St Thomas’ Hospital London. Exclusion criteria included a history of severe compromise at birth requiring prolonged resuscitation, a diagnosed chromosomal abnormality, any treatment for clinically significant brain injury, and any contraindication to MRI scanning. Participants with visible abnormality on MRI of possible/likely clinical significance following reporting by an experienced paediatric neuroradiologist were also excluded, including those with isolated cranial non-brain anomalies, major lesions within white matter, cortex, cerebellum, and/or basal ganglia, and microcephaly (head circumference < 1^st^ centile). Incidental findings with unlikely significance for clinical outcome where not excluded, such as birth related subdural haemorrhages, isolated subependymal cysts, and scattered punctate lesions.

At 18 months of corrected age, all participants were invited to complete neurodevelopmental follow-up assessments. After exclusion of participants without complete follow-up, demographic, or environmental data (detailed below), the final analysis samples comprised 121 participants (with a total of 185 baseline and follow-up scans) for the structural imaging analysis, and 110 participants (with 166 scans) for the functional imaging analysis. Sample characteristics of the final sample of 121 participants can be found in Table 1.

**Table 1.**
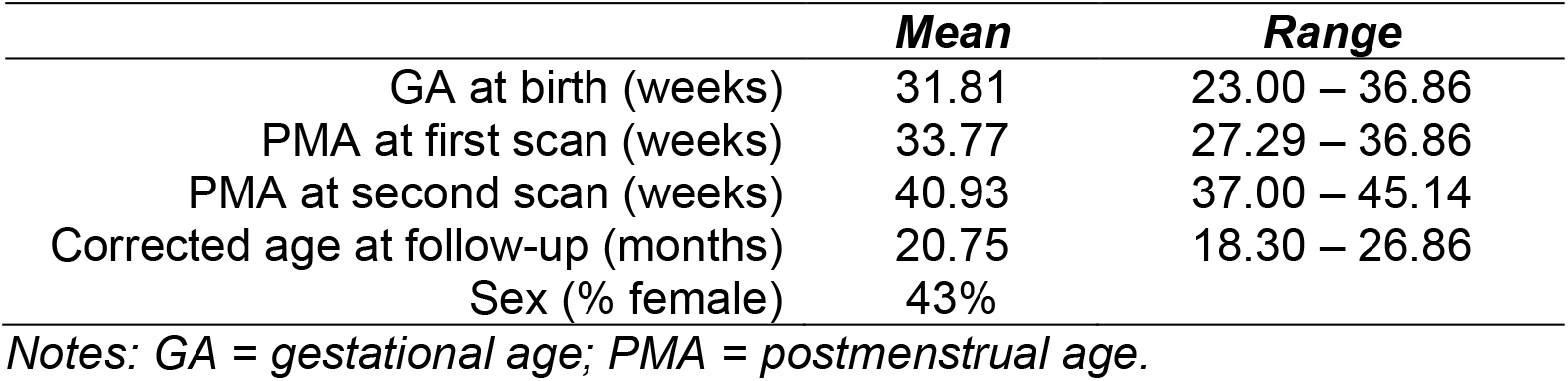
Sample characteristics of full analysis sample

### 2.2. Imaging acquisition

Infants underwent neuroimaging using a 3 T Philips Achieva system (Philips Medical Systems). All infants were scanned without sedation but during natural sleep in a scanner environment optimized for safe and comfortable neonatal imaging, including a dedicated transport system, positioning device and a customized 32-channel receive coil, with a custom-made acoustic hood (Hughes et al., 2017). All scans were supervised by a neonatal nurse and/or paediatrician who monitored heart rate, oxygen saturation and temperature throughout the scan.

T2-weighted images were obtained using a Turbo Spin Echo sequence, acquired in two stacks of 2D slices (in sagittal and axial planes), using parameters: TR = 12 s, TE = 156 ms, SENSE factor 2.11 (axial) and 2.58 (sagittal), acquisition resolution 0.8 × 0.8 × 1.6 mm^3^ with 0.8 mm slice overlap, reconstructed to 0.5 mm isotropic resolution.

High temporal resolution BOLD functional MRI optimized for neonates was acquired over 15 min 3 s (2300 volumes) using a multislice gradient-echo echo planar imaging (EPI) sequence with multiband excitation (multiband factor 9), using parameters: TR = 392 ms, TE = 38 ms, flip angle = 34°, acquisition resolution 2.15 mm isotropic.

### 2.3. Structural data pre-processing

Motion-correction and slice-to-volume reconstruction of T2 weighted images were carried out using a dedicated algorithm as previously described (Cordero-Grande et al., 2018). Each image was subsequently registered to a week-specific template (https://gin.g-node.org/BioMedIA/dhcp-volumetric-atlas-groupwise; constructed as described in Schuh et al., 2018), according to the individual’s postmenstrual age at scan, using the symmetric normalisation (SyN) algorithm from Advanced Normalisation Tools (ANTs), version 3.0 (Avants and Gee, 2004). Affine and non-linear transformations were performed, with non-linear transformations used to create deformation tensor fields in template space. The resulting tensor fields were used to calculate scalar Jacobian determinants, which were subject to logarithmic transformation, using ANTs. Log-Jacobian determinant maps (hitherto referred to as Jacobians) were then registered to a common space corresponding to the 40-week neonatal template from the extended dHCP volumetric atlas (Schuh et al., 2018). However, note that due to the derivation of the Jacobians from registrations to the individual week-specific templates (rather than to the 40-week template), these maps represent (head-size adjusted) regional volume *relative to the relevant age group* (i.e., the week-specific template). They can therefore be understood as volumetric deviations from the age-norm (i.e., a greater Jacobian reflects a greater regional volume relative to age-matched neonates at time of scan). Jacobian maps were smoothed with a 3 mm Gaussian smoothing kernel and masked with a cortical and subcortical grey matter mask defined in template space for further analysis.

### 2.4. Functional data pre-processing

Functional data were pre-processed using an in-house pipeline optimized for neonatal imaging (Fitzgibbon et al., 2020), including dynamic distortion and intra- and inter-volume motion correction for each participant. Motion, cardiorespiratory and multiband acquisition artefacts, as well as single-subject ICA noise components identified using FSL FIX (Salimi-Khorshidi et al., 2014) were regressed out. Images were subsequently registered into native T2 space using boundary-based registration and then non-linearly transformed to the dHCP 40-week template (Schuh et al., 2018). We excluded participants with high framewise displacement (FD > 1.5IQR + 75th centile) in more than 230 (10%) of the 2300 volumes. Images were smoothed with a 3 mm Gaussian smoothing kernel, bandpass filtered at 0.01-0.1Hz and masked with a cortical grey matter mask defined in template space (in contrast to the structural image analysis mask, this mask did not include subcortical or cerebellar deep grey matter, given that the bias towards improved signal-to-noise ratio on the cortical surface is magnified by multiband acceleration).

In order to derive voxelwise degree centrality maps (Fenn-Moltu et al., 2022), the timeseries from each voxel within the cortical grey matter mask (in template space) were extracted and a correlation matrix constructed from pairwise Pearson correlations between each pair of voxels. Correlations below 0.20 were excluded in order to avoid including spurious connections likely caused by noise. Weighted degree centrality was calculated for each voxel as the sum of correlations between this voxel and all other voxels within the mask. Thus, high degree centrality reflects that a voxel is highly connected to many other voxels. Voxelwise degree centrality maps were subsequently standardised to z-scores in order to allow for comparisons across participants. Thus, degree centrality values reflect centrality relative to the rest of the brain rather than absolute weighted degree centrality.

### 2.5. Behavioural outcome data

The parent-rated Child Behavior Checklist 1 1/2 - 5 (CBCL; Achenbach and Rescorla, 2000) was used to assess infant behavioural problems and psychopathology. Subscales derived from the CBCL for further analysis were the *Emotional reactivity, Anxiety/Depression, Somatic, Withdrawn, Sleep, Attention*, and *Aggressive* subscales. The Bayley Scale of Infant Development, Third Edition (Bayley-III; Bayley, 2006) was used in order to assess psychomotor development, yielding age-adjusted *Cognitive, Motor*, and *Language* composite scores.

### 2.6. Socio-demographic data

Parental postcode at the time of infant birth was used to derive an Index of Multiple Deprivation (IMD) score (Department for Communities and Local Government, 2011; https://tools.npeu.ox.ac.uk/imd/) as a measure of socioeconomic status. IMD is based on seven domains of deprivation within each neighbourhood: income, employment, education, skills and training, health and disability, barriers to housing and services, and living environment and crime. Higher IMD values indicate higher deprivation.

Parents completed a questionnaire adapted from the Cognitively Stimulating Parenting Scale reported in the study by Wolke et al. (2013). It consists of 28 items adapted in the Home Observation for Measurement of the Environment Inventory (Caldwell and Bradley, 1984), and assesses the availability and variety of experiences that promote cognitive stimulation in the home (for more details, see Supplemental Materials and Bonthrone et al., 2020).

### 2.7. Statistical analysis

#### 2.7.1. Principal Component Analysis on behavioural outcomes

Subscales from the CBCL and Bayley-III were subjected to Principal Component Analysis (PCA) in order to derive meaningful components of covarying psychopathology and psychomotor development. CBCL subscales were regressed against corrected age at assessment, and residuals from these regressions were subsequently used. All variables were scaled to a mean of 0 and unit variance before PCA was performed. A permutation testing procedure was used to identify significant components, as described in more detail elsewhere (Vanes et al., 2021). To assess meaningful contribution of individual variables to the identified PCs, we defined the loading threshold as 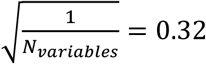, equivalent to the loading value if all 10 variables included in the PCA loaded equally onto the same component. Loadings greater than this value were considered to be meaningful. Two resulting behavioural principal components (PC1 and PC2; see results for details) were used in subsequent imaging and socio-demographic analyses.

#### 2.7.2. Longitudinal brain development and behaviour

We assessed neonatal longitudinal development of brain structure and function, and their respective relationships with behavioural outcomes at 18 months, using nonparametric mixed-effects marginal models with the FSL Sandwich Estimator (FSL-SwE) tool (Guillaume et al., 2014). Separate models were fit for brain structure (voxelwise log-Jacobian volumes) and function (voxelwise degree centrality). In order to robustly differentiate between cross-sectional and longitudinal age effects, we deconstructed postmenstrual age (PMA) at scan into its cross-sectional and longitudinal components (Neuhaus and Kalbfleisch, 1998). Cross-sectional PMA was each participant’s mean PMA across timepoints centred with respect to the whole sample 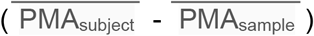, henceforth referred to as *Age*. Longitudinal PMA was the within-subject centred PMA at each timepoint 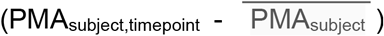, henceforth referred to as *Time*, as it represents the within-subject change over time while taking into account the temporal distance between scans.

Each of the two fitted models included all participants’ relevant voxelwise maps (log-Jacobian or degree centrality, respectively) as dependent variable. Independent variables for both models were GA at birth, sex, IMD, Stimulating Parenting Scale, both behavioural principal components (PC1 and PC2), Age, Time, and interactions between each behavioural component and Time. The structural imaging model also included as covariate a continuous variable indicating the temporal distance (in days) from the weekly template that each scan was registered to. The functional imaging model also controlled for in-scanner head motion during the resting-state acquisition (total number of framewise displacement outliers).

Effects of interest were the main effects of PC1, PC2, and their respective interactions with Time. This model therefore allowed for an investigation of the association between average (structural or functional) brain metrics and behavioural outcome, as well as between longitudinal change in brain metrics and behavioural outcome (controlling for GA at birth, sex, IMD, Stimulating Parenting, and Age). Statistical inferences for effects of interest were achieved using a non-parametric Wild Bootstrap procedure with 999 iterations (Guillaume and Nichols, 2015) with threshold free cluster enhancement (TFCE) (Smith and Nichols, 2009).

#### 2.7.3. Socio-demographic factors and behaviour

To assess relationships between 18-month behavioural outcomes and non-imaging socio-demographic (clinical, demographic and environmental) variables, we constructed two linear regression models, regressing each behavioural principal component, respectively, against gestational age (GA) at birth, corrected age at follow-up assessment, sex, IMD, Cognitively Stimulating Parenting score, and the remaining behavioural principal component.

## 3. Results

### 3.1. Principal Components of infant behavioural outcome

PCA on CBCL and Bayley-III subscales with non-parametric permutation testing yielded two significant principal components (PC1 and PC2, see Figure 1). PC1 showed meaningful loadings (>0.32) for the emotional reactivity, anxiety/depression, withdrawn, attention, and aggressive subscales of the CBCL, and is therefore termed the “Psychopathology” component. Higher PC1 scores indicate increased behavioural problems as reflected in these measures. PC2 showed meaningful loadings (>0.32) for all Bayley-III subscales and is therefore termed the “Psychomotor functioning” component. Higher PC2 scores indicate better cognitive, language, and motor development. Unthresholded loadings for both components can be found in Supplementary Figure S1.

**Figure 1.**
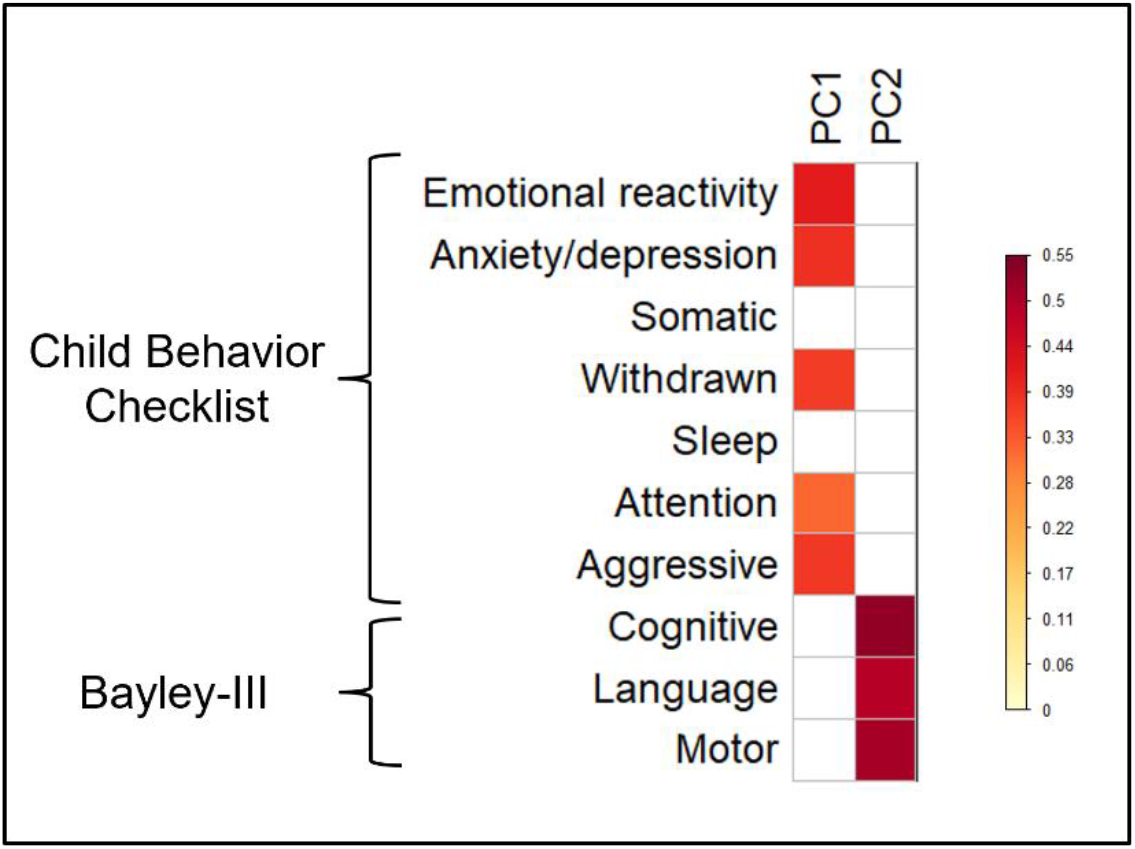
Heatmap of loadings of each variable on PC1 (Psychopathology) and PC2 (Psychomotor functioning), thresholded at 0.32.

### 3.3. Neonatal structural brain development and behavioural outcomes

PC2 (psychomotor functioning at 18 months) was significantly positively related to log-Jacobian values in a large cluster in the right cerebellum (6613 voxels, TFCE-corrected *p* = .014). As seen in Figure 2, this indicates that greater neonatal whole-brain adjusted right cerebellar volume (relative to other neonates who underwent MRI at the same age) is associated with better psychomotor functioning at 18 months, after controlling for GA at birth, sex, IMD, SPS, Age, Time, and PC1.

**Figure 2.**
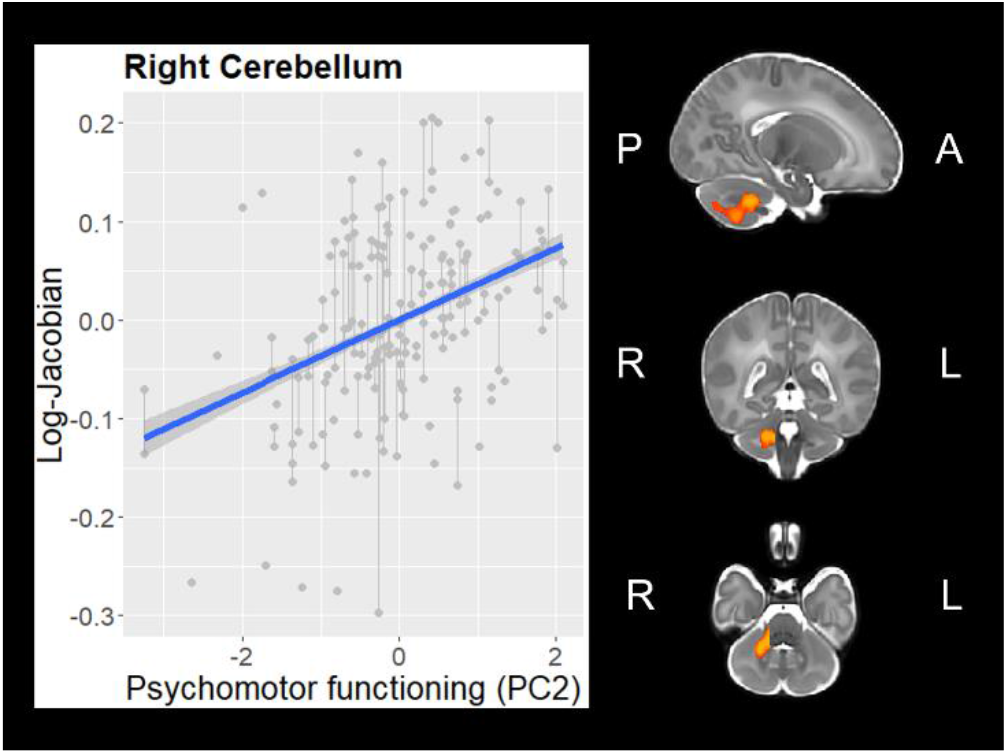
Association between Psychomotor functioning (PC2) and right cerebellar regional volume (controlling for gestational age at birth, sex, age at scan, time, and PC1)

Furthermore, there was a significant PC1 × Time interaction in two subcortical clusters: right thalamus (6287voxels, TFCE-corrected *p* = .008) and bilateral subthalamic nuclei (STN; 7271 voxels, TFCE-corrected *p* = .007). As displayed in Figure 3, regional volume in both clusters showed a relative reduction over time. Note that this does not reflect an absolute reduction of subcortical volumes, but rather reflects that the volume of these structures increasingly negatively deviates from the age-appropriate template as term-equivalent age approaches. Importantly, this deviation over time was moderated by PC1, indicating that the comparative reduction in regional volume was accelerated in infants with higher Psychopathology at 18 months.

**Figure 3.**
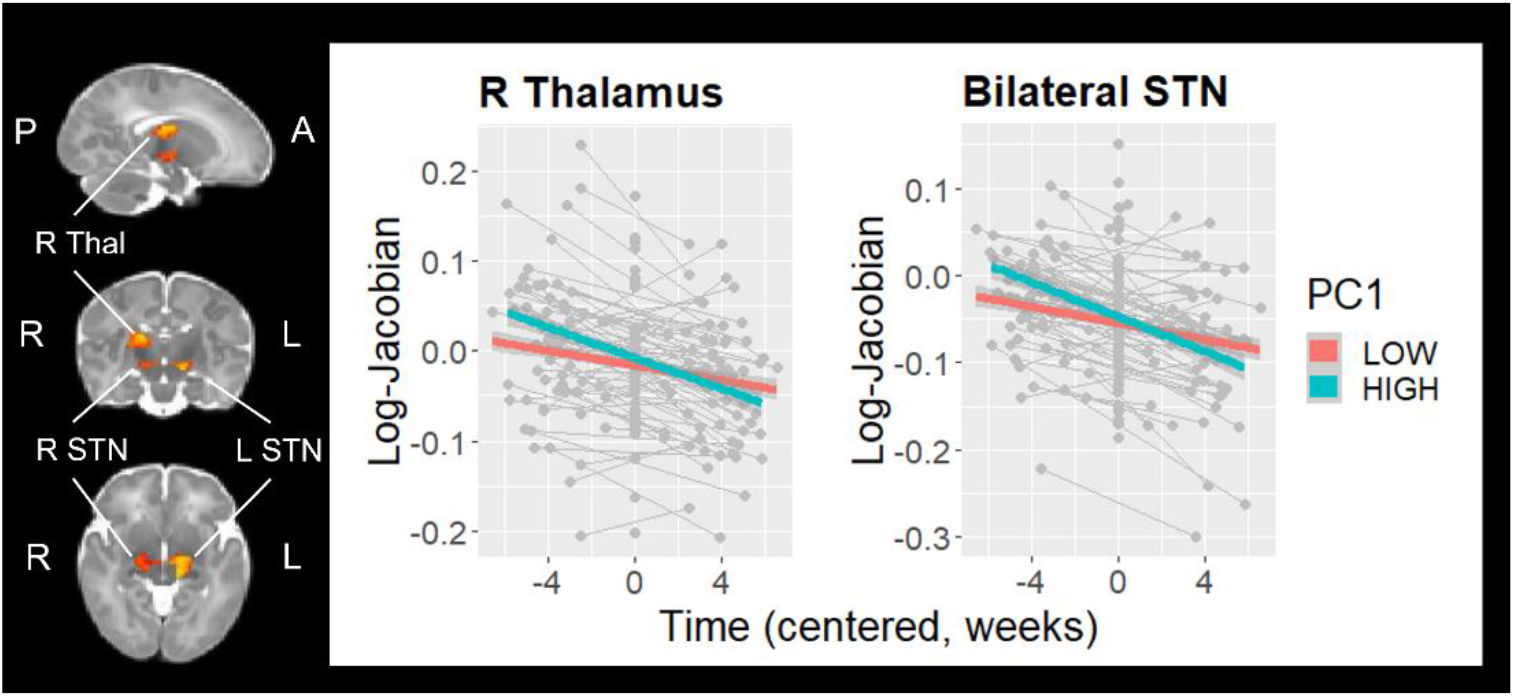
Psychopathology (PC1) × Time interaction on relative regional volumes of right thalamus and bilateral subthalamic nuclei (STN), controlling for gestational age at birth, sex, age at scan, time, and PC2. For visualisation purposes only, PC1 is depicted as a categorical variable following a median split.

### 3.4. Neonatal functional brain development and behavioural outcomes

There was a significant PC2 × Time interaction in several clusters of functional degree centrality, encompassing predominantly left sensorimotor cortex (381 voxels, TFCE-corrected *p* = .004) and posterior cingulate cortex (98 voxels, TFCE-corrected *p* = .013), indicating that a more rapid reduction in relative degree centrality in these regions occurred in infants with better psychomotor functioning at 18 months (Figure 4). There were no significant main effects of PC1, PC2, or PC1 × Time interaction for degree centrality.

**Figure 4.**
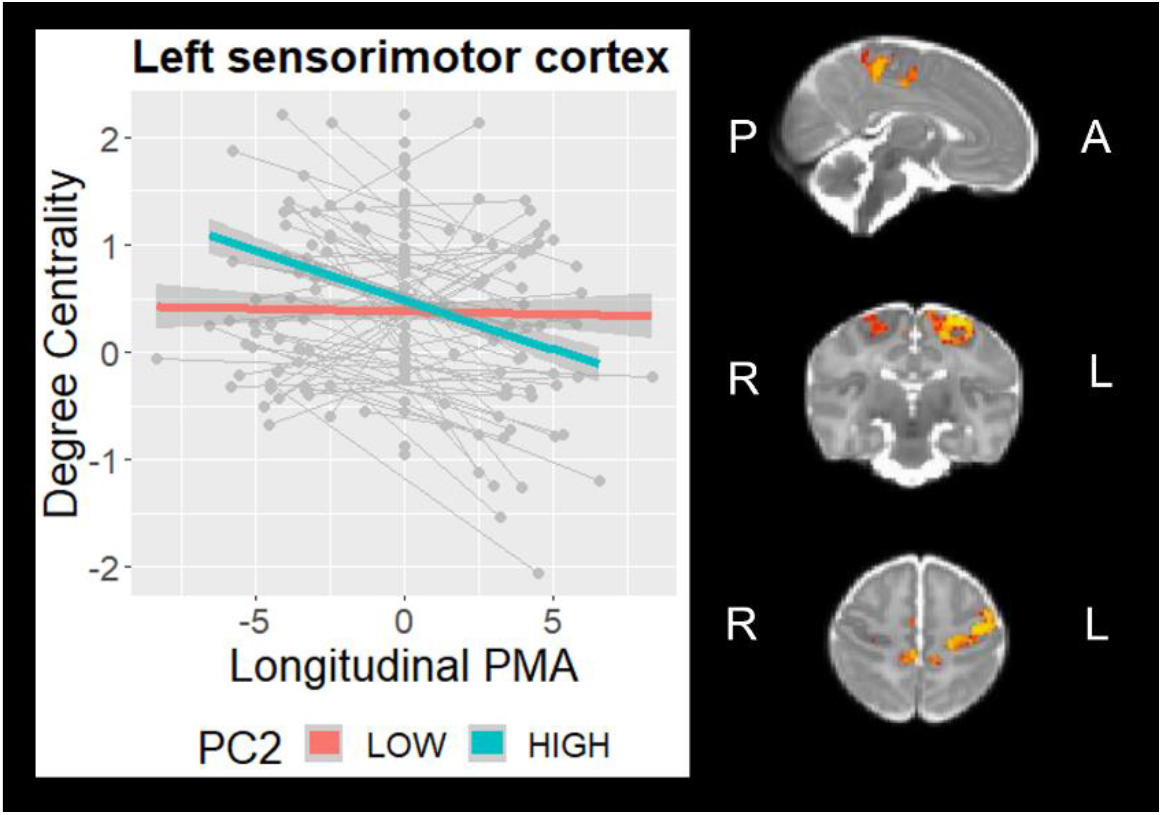
Psychomotor functioning (PC2) × Time interaction on degree centrality in sensorimotor cortex (controlling for gestational age at birth, sex, age at scan, time, and PC2). For visualisation purposes only, PC2 is depicted as a categorical variable following a median split.

### 3.2. Socio-demographic associations with behavioural outcomes

We conducted two linear regression analyses regressing PC1 (or PC2) on GA at birth, age at assessment, sex, IMD, Cognitively Stimulating Parenting Scale scores and PC2 (or PC1). There was a significant positive effect of IMD (beta = 0.04, *p* = .006) and negative effect of Cognitively Stimulating Parenting Scale scores (beta = -0.14, *p* = .003) on PC1, indicating that higher socio-economic deprivation and lower cognitively stimulating parenting are independently associated with increased psychopathology at 18 months (Table 2; Figure 5A). In contrast, for PC2, there were significant effects of GA at birth (beta = 0.10, *p* = .008) and IMD (beta = -0.03, *p* = .006), indicating that higher gestational age at birth and greater socio-economic advantage predicted better psychomotor functioning at 18 months (Table 2; Figure 5B).

**Table 2.** Regression analysis summary for socio-demographic variables predicting behavioural outcomes

|  | Estimate | Standard error | p-value |
| --- | --- | --- | --- |
| **Dependent variable: PC1** |  |  |  |
| GA at birth | -0.09 | 0.05 | .066 |
| Age at assessment | 0.05 | 0.11 | .677 |
| Sex | -0.36 | 0.31 | .243 |
| IMD | 0.04 | 0.01 | **.006** |
| Stimulating Parenting Scale | -0.14 | 0.05 | **.003** |
| PC2 | 0.24 | 0.12 | **.043** |
| **Dependent variable: PC2** |  |  |  |
| GA at birth | 0.10 | 0.04 | **.008** |
| Age at assessment | -0.10 | 0.08 | .228 |
| Sex | 0.30 | 0.24 | .210 |
| IMD | -0.03 | 0.01 | **.005** |
| Stimulating Parenting Scale | 0.03 | 0.04 | .449 |
| PC1 | 0.15 | 0.07 | **.043** |

Notes: GA = gestational age; IMD = Index of Multiple Deprivation

**Figure 5.**
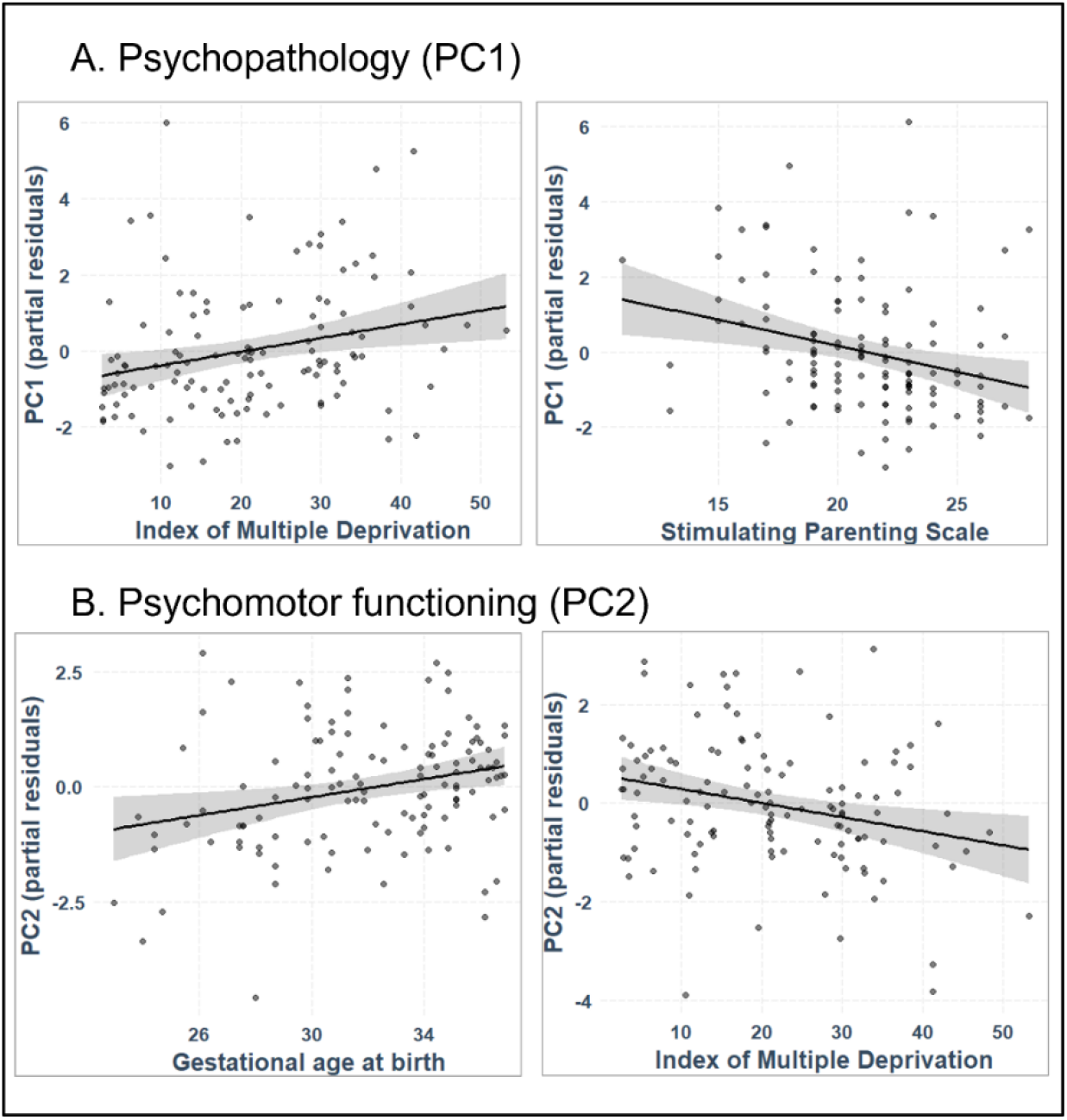
Partial regression scatterplots for significant effects of Index of Multiple Deprivation (IMD) and Stimulating Parenting Scale (SPS) on PC1 (A), and of IMD and gestational age (GA) on PC2 (B). Both regression models included GA at birth, sex, age at assessment, IMD, and SPS as independent variables, and points depict partial residuals.

## 4. Discussion

We investigated longitudinal structural and functional brain development and its association with behavioural outcomes in a large sample of infants born preterm. By including independent measures on both psychopathology and psychomotor functioning within the same models, we were able to tease apart the distinct neural correlates of these two developmental domains while controlling for key socio-demographic factors known to influence both brain development and behaviour. We also explored the direct relationships between socio-demographic variables and toddler outcomes in this preterm cohort.

We used PCA to derive latent dimensions of behavioural development at 18 months of age. Two significant components emerged, reflecting orthogonal developmental outcomes related to psychopathology and psychomotor functioning, respectively. PC1, driven by all but the more physical (somatic and sleep related) subscales of the CBCL, represents an increased burden of early psychopathology, while PC2 encapsulates performance on all three (cognitive, language, and motor) Bayley-III subscales. It is worth highlighting the conceptual distinctiveness of these two components, as reflected in their unthresholded component loadings. These show that in the case of PC1, increased psychopathology covaries with poorer psychomotor performance (consistent with generally less favourable outcomes). In contrast, along PC2, better psychomotor performance covaries with subthreshold increased, rather than decreased, psychopathology (thus possibly reflecting more “pure” psychomotor functioning). This pattern is largely consistent with our previous observations in an independent cohort of slightly older very preterm born children (Vanes et al., 2021). Including the two components jointly in subsequent analyses allowed us to assess the neural and socio-demographic correlates of one while controlling for the other.

At the structural brain level, we made two observations: first, neonatal right cerebellar volume across both time points was related to PC2 (psychomotor functioning), while rate of change in neonatal subcortical volume was related to PC1 (psychopathology). We evaluated neonatal regional brain volume by deriving Jacobian determinant maps from nonlinear registrations to week-specific average templates – voxelwise values thus reflecting the deviation in volume from an average brain at the respective postmenstrual age at scan. Note that due to the inclusion of a range of gestational ages in the weekly templates (Schuh et al., 2018), more term-born infants are used to construct templates at older scan ages. It is therefore expected that our sample of preterm-born infants would show greater negative deviations from the respective templates as term-equivalent age approaches, given that preterm infants are known to show widespread reductions in regional brain volume (Alexander et al., 2019; Dimitrova et al., 2021; Peterson et al., 2003). This becomes particularly relevant when interpreting the second finding.

Our first finding indicates that across both MRI timepoints (preterm birth and term-equivalent age), smaller than average right cerebellar volume is associated with poorer psychomotor performance at 18 months of age; an observation highly consistent with the existing literature. The role of the cerebellum in motor coordination and, more recently cognition, is widely acknowledged (Buckner, 2013), with aspects of motor, language, and cognitive development thought to be closely intertwined and dependent on cerebellar function (Diamond, 2000; Mariën and Borgatti, 2018). The cerebellum is particularly vulnerable to injury following preterm birth (Messerschmidt et al., 2005; Volpe, 2009), and disrupted cerebellar development related to perinatal brain injury is associated with poorer neuromotor and mental outcomes in toddlerhood (Bolduc and Limperopoulos, 2009; Messerschmidt et al., 2008). Even controlling for overt injury (Van Kooij et al., 2012) and other perinatal factors (Cheong et al., 2016), cerebellar volume at term-equivalent age has been shown to be associated with neurodevelopmental outcome following preterm birth. A recent longitudinal study showed that smaller neonatal and childhood cerebellar volumes, as well as slower growth rate, were related to poorer IQ, language, and motor function at 7 years of age (Matthews et al., 2018). Our findings confirm the specific association between neonatal cerebellar volume and neurodevelopmental outcome, and localise the effect more specifically to right cerebellum; however, cerebellar growth in the first weeks after birth does not appear to be sensitive to individual differences in psychomotor functioning in toddlerhood.

In contrast, psychopathology outcomes (PC1) in toddlerhood were related to longitudinal change in neonatal subcortical brain volume across the first weeks after birth. We found that relative volumes of the subthalamic nuclei and right thalamus showed a reduction over time from birth to term-equivalent age across the cohort – reflecting that volumes increasingly deviated (negatively) from the weekly template as term-equivalent age approaches. Crucially, this effect was most pronounced in those infants showing increased psychopathology at 18 months. This suggests that infants who show slower regional growth (relative to the average brain) in these regions are those most likely to experience increased behavioural and emotional problems in toddlerhood, after controlling for psychomotor and socio-demographic factors.

Smaller basal ganglia and thalamus volumes have been previously reported in preterm-born neonates (Boardman et al., 2006; Makropoulos et al., 2016; Srinivasan et al., 2007) and children (Kesler et al., 2004; Peterson et al., 2000) compared to their term-born peers, with likely implications for developmental outcome given the role of these structures in a wide range of cognitive, affective, and somatosensory functions (Arsalidou et al., 2013). Interestingly, whereas previous studies have predominantly found associations between thalamic and basal ganglia volumes and cognitive and motor outcomes in preterm children (Loh et al., 2017) and adults (Nosarti et al., 2014), our findings suggest that when specifically taking psychomotor functioning into account, early growth rate in these regions appear to be more relevant to psychopathology outcomes. Prominent involvement of the subthalamic nuclei is of particular interest here. Within the basal ganglia circuitry, the role of subthalamic nucleus in movement regulation has long been well documented (Hamani et al., 2004); however, there is also an increasing recognition of its function within the limbic system (Haegelen et al., 2009; Mavridis et al., 2013). Integrated into a limbic subcortical circuit and as the only structure within the basal ganglia to receive direct input from motor, premotor, and medial prefrontal areas (Chudasama et al., 2003), it has been suggested that the subthalamic nucleus is a key relay station for emotional and associative information, for instance integrating affective and motivational information into movement realisation (Haegelen et al., 2009). Disruption of the subthalamic nucleus can therefore have widespread implications for behaviour, consistent with affective symptoms (such as apathy and depression) frequently emerging after deep brain stimulation of this structure in Parkinson’s disease (Castrioto et al., 2014; Wang et al., 2018). Our findings point towards early growth rate of subthalamic nuclei and right thalamus following preterm birth as a potential marker of risk for behavioural and affective problems in early development.

We also investigated functional connectivity of the neonatal preterm brain and its associations with 18-month outcomes. Adult-like topography of functional resting-state networks can already be observed in the neonatal period (Eyre et al., 2021), with a maturational sequence progressing from primary to higher order cortical networks in infancy (Gao et al., 2017). Degree centrality is a useful metric quantifying the strength of connectivity of each voxel with the rest of the cortex, allowing for an evaluation of the importance of each region in the whole-brain network. Research in term-born neonates using this metric has found the most connected (“hub”) regions in infancy to be in motor and primary sensory cortices (Fransson et al., 2011). While strength and heterogeneity of functional connectivity increases with age in most networks during infancy (Cao et al., 2017), recent work in predominantly term-born neonates of the dHCP cohort has highlighted that sensorimotor cortex shows a decrease in relative degree centrality with increasing postmenstrual age (Fenn-Moltu et al., 2022). This may reflect a normative fine-tuning in the regional specificity of sensorimotor connectivity with increasing exposure to the surrounding environment, consistent with the emergence of functional specialisation of network connectivity in this period (Cao et al., 2017). Intriguingly, in the current study we found that a more rapid decrease in relative degree centrality in sensorimotor cortices was associated with better psychomotor functioning at 18 months of age in preterm infants. This finding suggests that a slower rate in the normative fine-tuning process of sensorimotor cortices during the neonatal period may be a risk factor for poorer cognitive, language, and motor skills in later years.

The brain-behaviour mapping undertaken in this study controlled for a range of relevant demographic, familial, and socio-economic factors known to play a role in child development. However, the direct relationships between these factors and behavioural outcomes themselves are also of substantial interest (Anderson, 2019). We found that area-level (social deprivation) and individual-level (cognitively stimulating parenting) environmental factors were differentially associated with psychopathology and psychomotor functioning. Consistent with a range of previous findings, higher socio-economic deprivation was associated both with increased psychopathology and poorer psychomotor functioning in toddlerhood. Studies have suggested that socio-economic status is an independent risk factor for adverse cognitive (Potijk et al., 2013; Wong and Edwards, 2013) and behavioural (Potijk et al., 2015) outcomes following preterm birth, whereby its impact adds to separate adverse effects of younger gestational age. Our results confirm this observation for psychomotor functioning, which was independently associated with gestational age at birth and socio-economic deprivation. In contrast, our results show that socio-economic deprivation, but not gestational age at birth, was predictive of psychopathology at 18 months after controlling for psychomotor functioning. Furthermore, a favourable home environment – specifically, more cognitively stimulating parenting – was selectively related to reduced psychopathology (but not better psychomotor functioning) in toddlerhood. This is consistent with our previous observations in preschool-aged children born very preterm (Vanes et al., 2021), and highlights the importance of the immediate family environment for behavioural and emotional development following preterm birth. Our findings demonstrate that environmental factors known to impact childhood outcomes following preterm birth can influence toddlers’ development in the first 18 months of life, underscoring the importance of targeting environmental interventions at the earliest stages of development (Anderson and Cacola, 2017).

An important strength of our study is the inclusion of longitudinal imaging in a substantial subset of preterm infants, allowing us to detect within-subject changes in brain structure and function related to outcome. It is possible that, given the wide range of gestational ages at birth in our sample (with a skew towards moderately preterm birth), the findings observed here extend to term-born infants. However, due to the lack of corresponding longitudinal imaging data in term-born infants studied as part of the dHCP cohort, we were unable to test this explicitly. Nevertheless, in light of the heightened neurodevelopmental risk with lower gestational age at birth, studying the neural and environmental mechanisms underlying adverse behavioural outcomes is particularly important in preterm cohorts specifically. Future research can further investigate structure-function relationships relative to neurodevelopmental processes underlying psychomotor, emotional, and behavioural functioning, while investigating how these insights might be leveraged to inform the nature and timing of early interventions to foster resilience in vulnerable infants.

## Data Availability

Data are available via http://www.developingconnectome.org/data-release/third-data-release/.

## Acknowledgments

This work was supported by the European Research Council under the European Union’s Seventh Framework Programme (FP7/20072013)/ERC grant agreement no. 319456 (dHCP project). The authors acknowledge infrastructure support from the National Institute for Health Research (NIHR) Mental Health Biomedical Research Centre (BRC) at South London, Maudsley NHS Foundation Trust and Institute of Psychiatry, Psychology and Neuroscience, King’s College London and the NIHR-BRC at Guys and St Thomas’ Hospitals NHS Foundation Trust (GSTFT). The authors also acknowledge support in part from the Wellcome Engineering and Physical Sciences Research Council (EPSRC) Centre for Medical Engineering at Kings College London [WT 203148/Z/16/Z], and the Department of Health through an NIHR Comprehensive Biomedical Research Centre Award (to Guy’s and St. Thomas’ National Health Service (NHS) Foundation Trust in partnership with King’s College London and King’s College Hospital NHS Foundation Trust). LDV is supported by a King’s Prize Fellowship [Wellcome Trust Institutional Strategic Support Fund; 204823/Z/16/Z] and the Medical Research Council [MR/S026460/1]. SFM is supported by a grant from the UK Medical Research Council [MR/N013700/1]. L.C-G. LCG is supported by a Beatriz Galindo Fellowship jointly funded by the Ministerio de Educación, Cultura y Deporte and the Universidad Politécnica de Madrid [BEAGAL18/00158]. TA and ADE received support from the Medical Research Council Centre for Neurodevelopmental Disorders, King’s College London [MR/N026063/1]. TA is supported by a MRC Clinician Scientist Fellowship [MR/P008712/1] and Transition Support Award [MR/V036874/1]. DB received support from a Wellcome Trust Seed Award in Science [217316/Z/19/Z]. CN is supported by the Medical Research Council [MR/S026460/1]. The views expressed are those of the authors and not necessarily those of the NHS, the National Institute for Health Research or the Department of Health. The funders had no role in the design and conduct of the study; collection, management, analysis, and interpretation of the data; preparation, review, or approval of the manuscript; and decision to submit the manuscript for publication.

