## Supplementary Materials for "Longitudinal neonatal brain development and socio-demographic correlates of infant outcomes following preterm birth"

### Supplementary Material

#### Cognitively Stimulating Parenting scale

Parents completed a questionnaire adapted from the Cognitively Stimulating Parenting Scale reported in the study by Wolke et al. (2013). It consists of 28 items, of which 21 are included in the Home Observation for Measurement of the Environment Inventory (Caldwell and Bradley, 1984) and 7 are added here. The questionnaire assesses the availability and variety of experiences that promote cognitive stimulation in the home. It has previously been shown to have acceptable internal consistency (Cronbach α = 0.77) (Wolke et al., 2013).

Of the 21 original items, 16 binary items capture the child's access to stimulating objects such as educational toys (6 items), parental interactions such as teaching numbers or colours (7 items), and parental behaviours such as reading habits (3 items). One item ("access to cassette player") was adapted to reflect technological advances appropriate at time of testing ("access to cassette/CD/DVD player"). Five Likert-scale items assess the frequency of cognitively stimulating experiences such as family excursions (4 items) and number of books in the home (1 item). Seven additional binary questions were added assessing whether the child has access to "YouTube", "computers/iPads/iPhone", "learning apps such as Peak-a-boo, Peppa Pig or Fish School", "gross motor toys such as trains cars bikes to sit on and push along", "a child-size table", "toys that teach household tasks such as sweeping, ironing, washing or other daily activities", and whether parents follow current affairs. A total sum score was calculated from all items (minimum 0, maximum 46).

#### Principal Component Analysis

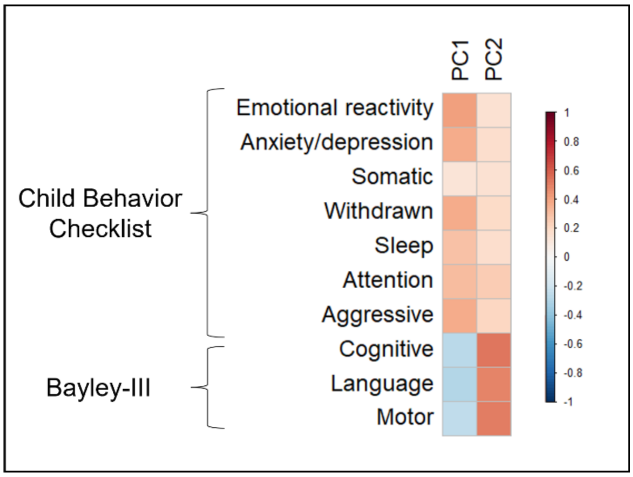


**Figure S1.** Heatmap of unthresholded loadings of each variable on PC1 (Psychopathology) and PC2 (Psychomotor functioning).
